# Acceptability of a virtual reality tool for early rehabilitation in intensive care units

**DOI:** 10.1101/2025.07.03.25330816

**Authors:** Florence Celant le Manac’h, Justine Saint-Aubert, Yoann Launey, Benoit Painvin, Anatole Lécuyer, Mélanie Cogné

## Abstract

**Introduction:** Virtual Reality (VR) is playing an increasingly important role in the medical field environment, whether for pain management or healthcare professionals’ training. The use of a VR device during a stay in Intensive Care Unit (ICU) could contribute to early rehabilitation’s management. Evaluation of the acceptability of such VR device on patients and healthcare professionals in these departments is the first step before considering integrating it, in order to ascertain the intention to use this device, but also to assess the possible obstacles to its use.

**Method:** We performed an acceptability study including patients and healthcare professionals from the ICUs of the University Hospital of Rennes, France. They answered a questionnaire based on the UTAUT 2 model, after a brief presentation of the VR device created to promote mental imagery early in ICU. Main judgment criterion was evaluating the acceptability via intention-to-use of this VR tool. Secondary objectives were to identify items influencing intention to use a VR tool in the healthcare professionals and patient populations.

**Results:** Sixty-eight healthcare professionals and fifty patients completed the questionnaire. We found a positive evaluation and a favorable intention to use the device in both populations. The main factors influencing intention to use were “perception of use” and “hedonic motivation” in both populations. Healthcare professionals attached importance to social influence, while patients prioritized “perceived ease of use”.

**Discussion:** Our results are in line with the literature on the a priori acceptability and intention to use virtual reality. However, this is the first study of virtual reality for early rehabilitation in ICU.

## Introduction

Mental imagery (i.e., observation and imagination of action) has already improved walking, balance and muscle tone in neurological populations (6-7). Mental imagery may allow us to provide early rehabilitation sessions in a population confined to Intensive Care Unit (ICU) with ICU-Acquired Weakness (ICU-AW, bilateral and symmetrical motor deficits predominantly in the proximal and lower limbs) that prevent them from performing real movements. Virtual reality (VR) has been shown to be helpful for active motor rehabilitation, for example after stroke (1–2), spinal cord injury (3), multiple sclerosis (4) or Parkinson’s disease (5) but could potentially be used to help mental imagery by simulating the action. The VERARE application (Virtual Environments for Rehabilitation After Reanimation) has been created by Inria Rennes, France, for allowing patients with ICU-AW to perform mental imagery of walking enhanced by VR in order to offer them early rehabilitation sessions *(please see Annex 1 for details concerning the VERARE application)*.

The main objective of our acceptability study is to evaluate the intention-to-use of the VR tool developed among a population of patients hospitalized in ICU and healthcare professionals working in ICU. The secondary objective of our study is to identify items influencing intention-to-use of a VR tool in the 2 populations studied. Our research hypotheses are that the intention to-use is positively influenced by: the perceived usefulness of the VR environment (H1), its perceived ease of use (H2), the social influence (H3), the facilitating conditions of use (H4), the hedonic motivation of use (H5), the adaptation to unit care (H6 for healthcare professionals), and the habits (H6 for patients, H7 for healthcare professionals).

## Methods

Thus, we performed a prospective, open-ended, monocentric acceptability study. Our main inclusion criterion for patients was to be hospitalized in one of the ICUs of the University Hospital of Rennes, France, for less than 3 months. For healthcare professionals, the main inclusion criterion was to have been working as one of the following professions (medical doctor, nurse, nursing auxiliary, physician or physiotherapist), in one of these units for more than one month. Exclusion criteria were for both populations: being no-French speaker, under legal protection, or deprived of liberty; having vigilance disorders, confusion or severe comprehension disorders preventing completion of the questionnaire; having a neurological central lesion.

We used 2 distinct questionnaires based on the UTAUT 2 acceptability model (“Unified theory of technology acceptance and use” which is the reference model used in the literature (8)) with both healthcare professionals and patients from September 2022 to May 2023. An e-mail was sent to healthcare professionals via a survey. Data were anonymized by the website. For all patients, the VR system was presented orally with the help of a support document *(please see Annex 1*) to illustrate the VR device (i.e., the VR headset computer, the cameras and various images of the VR environments created). Concerning ethics’ approvals, a declaration of accordance with the CNIL (“Commission Nationale de l’Informatique et des Libertés”) was made on August 20th, 2019 (declaration number: 2205295 v0). The study was submitted to the Ethics Committee of the University Hospital of Rennes, France, which issued a favorable opinion on 19/12/2020 (Opinion n° 20.161). An information letter was sent to introduce the study to the participants. An objection form on the last page of the information letter was also enclosed. Participants could return it within 3 weeks if they decided to withdraw from the study.

To test the reliability of the questionnaires, we first performed a Cronbach’s alpha test. Then we used non-parametric tests, such as correlation matrix with Spearman’s Rho and logistic regressions. Statistics were compiled using XLSTAT, JAMOVI and SAS, V9.4® (Statistical Analysis System). P<0.05 was considered significant. To compare both populations, we performed a logistic regression to look for factors associated with intention-to-use after deleting the questions which were not similar.

## Results

For including healthcare professionals, we sent e-mails to 200 of them, 71 answered, but 10 did not answer to our primary judgment criterion. Fifty-five patients were screened, 3 refused to participate and 1 subsequently withdrew their consent. Finally, we considered the results of 112 participants (61 healthcare professionals, 51 patients *(please see Table 1 for details concerning their general characteristics)*. The Cronbach’s alpha performed resulted 0,96 for both populations, which ensured the reliability of the questionnaires. The intention-to-use of the VR tool developed was quoted at 5.3/7 for patients and 6.0/7 for healthcare professionals and patients. Hypotheses H1 to H5 were supported in both populations. Hypothesis H6 was supported in the healthcare professional’s population only. Hypothesis H6 for patients and H7 for healthcare professionals were not supported *(Figure 1)*.

**Table 1:** General data of the 112 participants included.

|  | **<u>Healthcare</u>** | **<u>Patients</u>** | **<u>Participants</u>** | **<u>Participants</u>** |
| --- | --- | --- | --- | --- |

*Table 1: General data of the 112 participants included*
|  | **<u>professionals</u>**<br>**N=61** | **N=51** | **<u>favorable for</u>**<br>**<u>using VR in</u>**<br>**<u>ICU</u>**<br>**N=91** | **<u>unfavorable</u>**<br>**<u>for using VR</u>**<br>**<u>in ICU</u>**<br>**N=21** |
| --- | --- | --- | --- | --- |
| Gender (W/M/missing data) | 47/13/1 | 17/34/0 | 54/36/1 | 10/11/0 |
| Medical doctors/paramedical staff | 10/57 | - | - | - |
| Age (years old) (average ± SD) | 38.9 ± 8.9 | 53.9 ± 16.6 | 44.8 ± 15.6 | 52.0 ± 12.0 |
| Comfortable with new technologies (/7) (Median [Q1; Q3]) | 4.9 [4; 6] | 4.0 [3; 6] | 4.8 [4; 6] | 3.4 [2; 5] |
| Overall assessment of the VR tool (/7) (Median [Q1; Q3]) | 5.5 [5; 7] | 5.2 [4; 6] | 5.6 [5; 6] | 4.7 [4; 5,5] |
| Intention to use the VR tool (/7) (Median [Q1; Q3]) | 6.0 [3; 7] | 5.3 [4; 7] | 6.3 [6; 7] | 2.9 [1; 4] |

**Figure 1:**
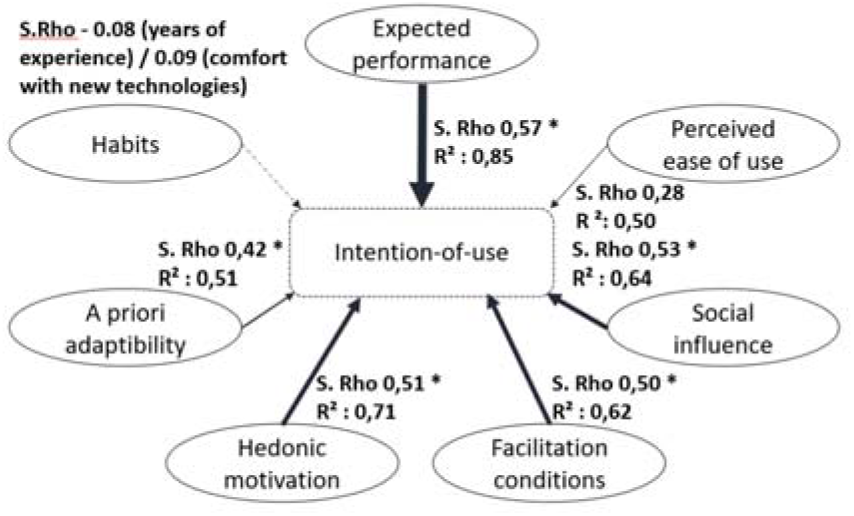
Verification of our hypotheses concerning factors influencing intention-to-use for healthcare professionals

**Figure 2:**
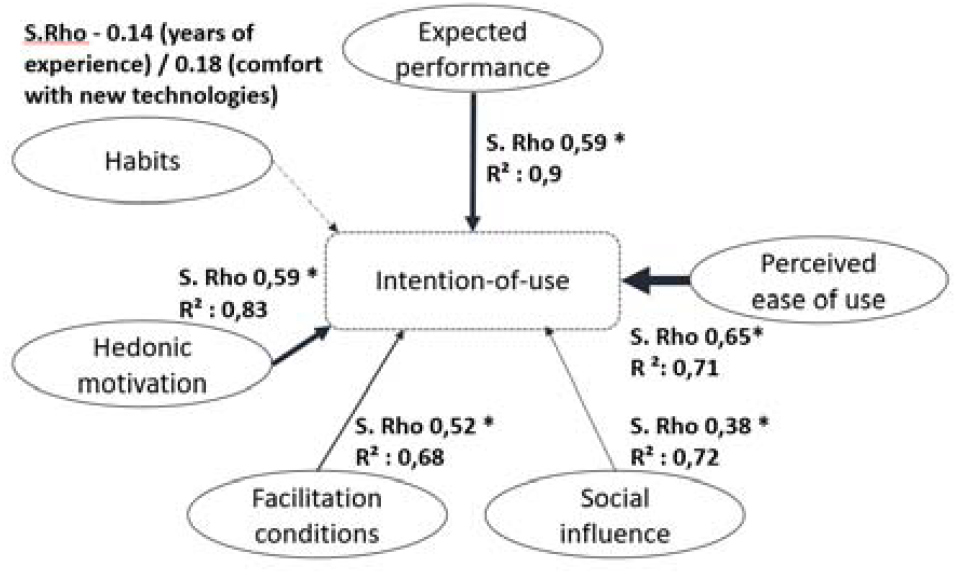
Verification of our hypotheses concerning factors influencing intention-to-use for patients

We described the average of each item contained in the acceptability questionnaire in Table 2.

**Table 2:**
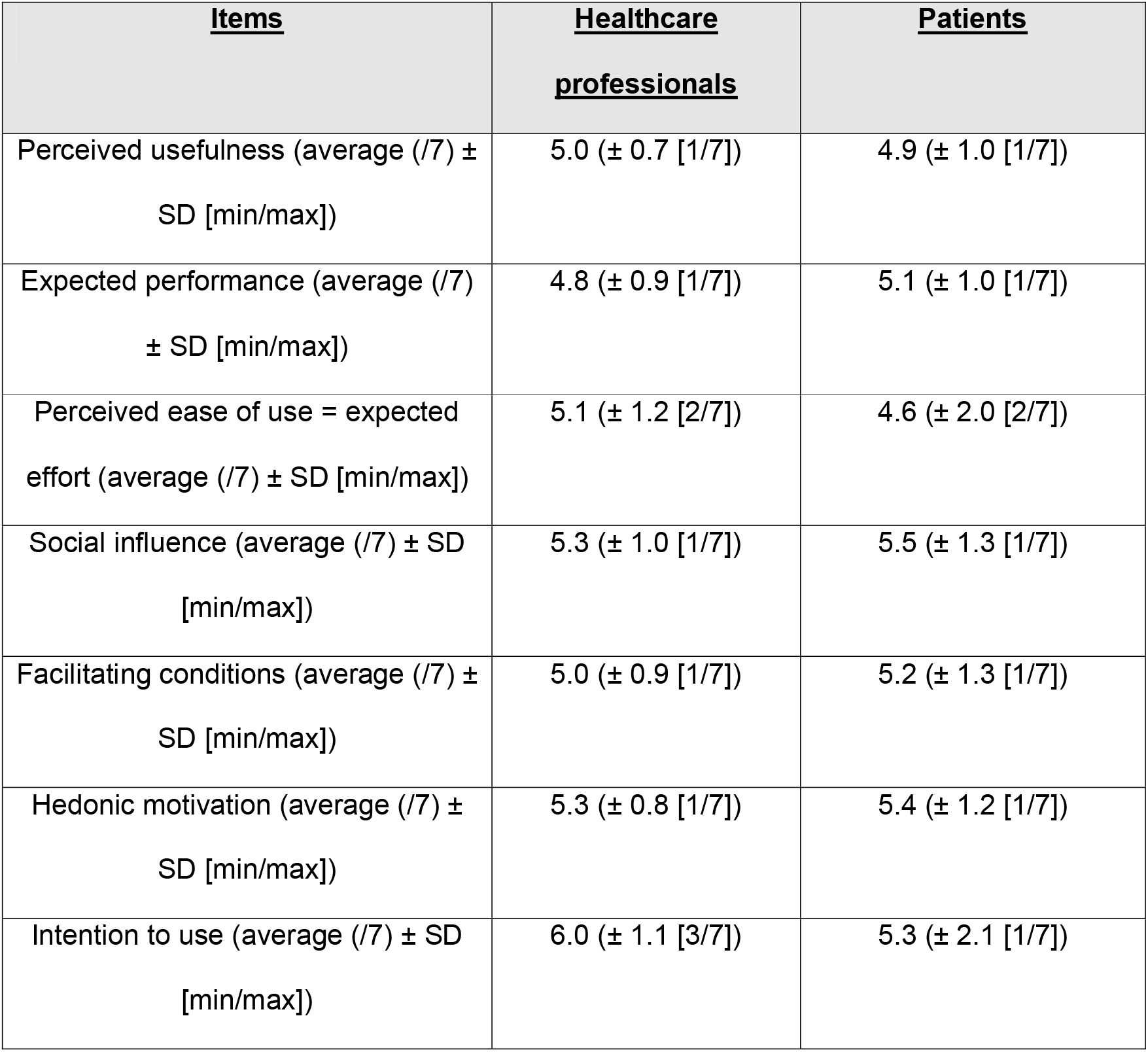
Descriptive analyses of answers of the 112 participants included.

| <u>Items</u> | <u>Healthcare professionals</u> | <u>Patients</u> |
| --- | --- | --- |
| Perceived usefulness (average (/7) ± SD [min/max]) | 5.0 (± 0.7 [1/7]) | 4.9 (± 1.0 [1/7]) |
| Expected performance (average (/7) ± SD [min/max]) | 4.8 (± 0.9 [1/7]) | 5.1 (± 1.0 [1/7]) |
| Perceived ease of use = expected effort (average (/7) ± SD [min/max]) | 5.1 (± 1.2 [2/7]) | 4.6 (± 2.0 [2/7]) |
| Social influence (average (/7) ± SD [min/max]) | 5.3 (± 1.0 [1/7]) | 5.5 (± 1.3 [1/7]) |
| Facilitating conditions (average (/7) ± SD [min/max]) | 5.0 (± 0.9 [1/7]) | 5.2 (± 1.3 [1/7]) |
| Hedonic motivation (average (/7) ± SD [min/max]) | 5.3 (± 0.8 [1/7]) | 5.4 (± 1.2 [1/7]) |
| Intention to use (average (/7) ± SD [min/max]) | 6.0 (± 1.1 [3/7]) | 5.3 (± 2.1 [1/7]) |

## Discussion

This acceptability study shows that the VERARE device developed is well accepted by patients and healthcare professionals in ICU. Furthermore, 79% of healthcare professionals and 72% of patients said they would be willing to use it in these units. The items that seemed to have the greatest influence on the intention to use in the healthcare professional’s population were “perceived usefulness” and “social influence”. It seems important to create a low-cost device (for a best use of public funds) to make them acceptable to the hierarchy of these services (“social influence”). We also found a significant correlation between “facilitation conditions”, “hedonic motivation”, “perceived ease of use” and “intention to use” in the healthcare professional’s population. The items that seemed to have the greatest influence on the intention to use in the patients’ population were in decreasing order: first “perceived ease of use”, then “hedonic motivation” and “expected performance”. Finally, “facilitation conditions” and “social influence” had an influence on intention-of-use in our study. We note that subjects unfavorable to the use of the device showed a lower comfort level with new technologies (Table 1). Our results show the importance of creating a device that is easy to use and fun to play with. Finally, despite different objectives, it appears that it would seem that, to encourage the use of a tool, it must be seen as useful to the person using it and compatible with their social environment. To the best of our knowledge there is no study of the acceptability of virtual reality as a tool for early rehabilitation in ICU. Our study provides an initial approach to assess the intention to use in these conditions. A potential limitation of our study is that they were different conditions concerning the completion of the questionnaires between our 2 populations. Thus, patients (interviewed individually) may have been positively influenced by the presence of an examiner, while healthcare professionals (interviewed by e-mail) were not. There was also a selection bias in our healthcare professional’s recruitment (34% of the healthcare professionals contacted via e-mail answered). Moreover, participants did not have the opportunity to test the virtual reality device in this acceptability study. The aim of the VERARE VR application presented will next to be tested to generate mental imagery of walking.

## Acknowledgements

the authors thank the patients and the therapists who answered the questionnaires

## Data Availability

on demand

## Annex 1. Presentation of the VERARE application

1A. General explanation concerning the VR tool: the acceptability survey concerned the acceptability of a VR application designed to improve walking in ICU. The VR application includes the use of a computer software (VERARE), a “HTC Vive Pro” virtual reality headset, and “HTC Vive base-stations”. The device can be taken to the patient’s bed in ICUs. The VERARE environment immerses the patient in a virtual world with a first-person projection.

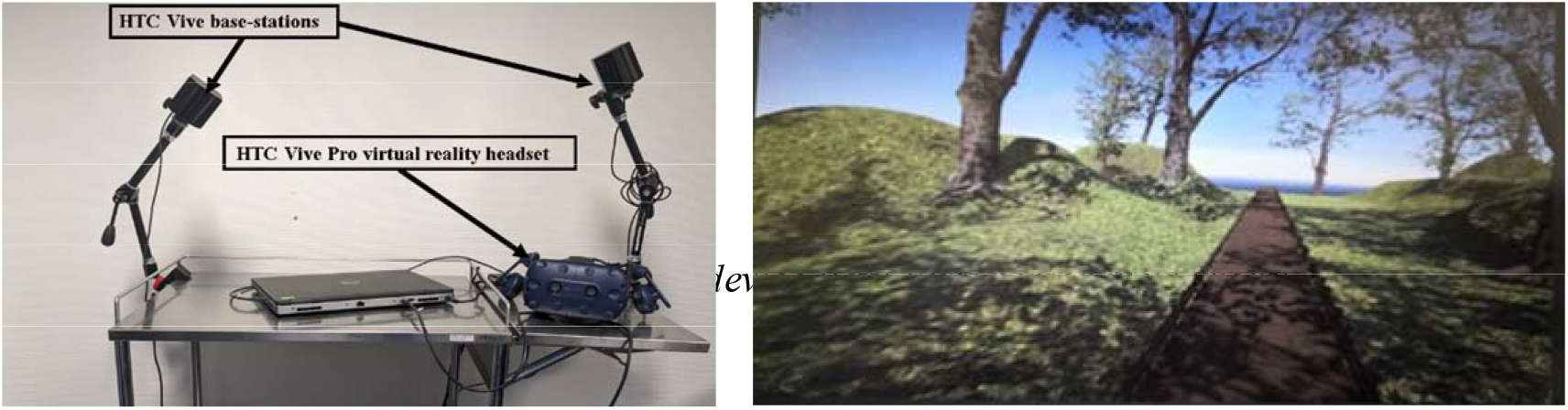

1B. Pictures presented to the participants of the acceptability study

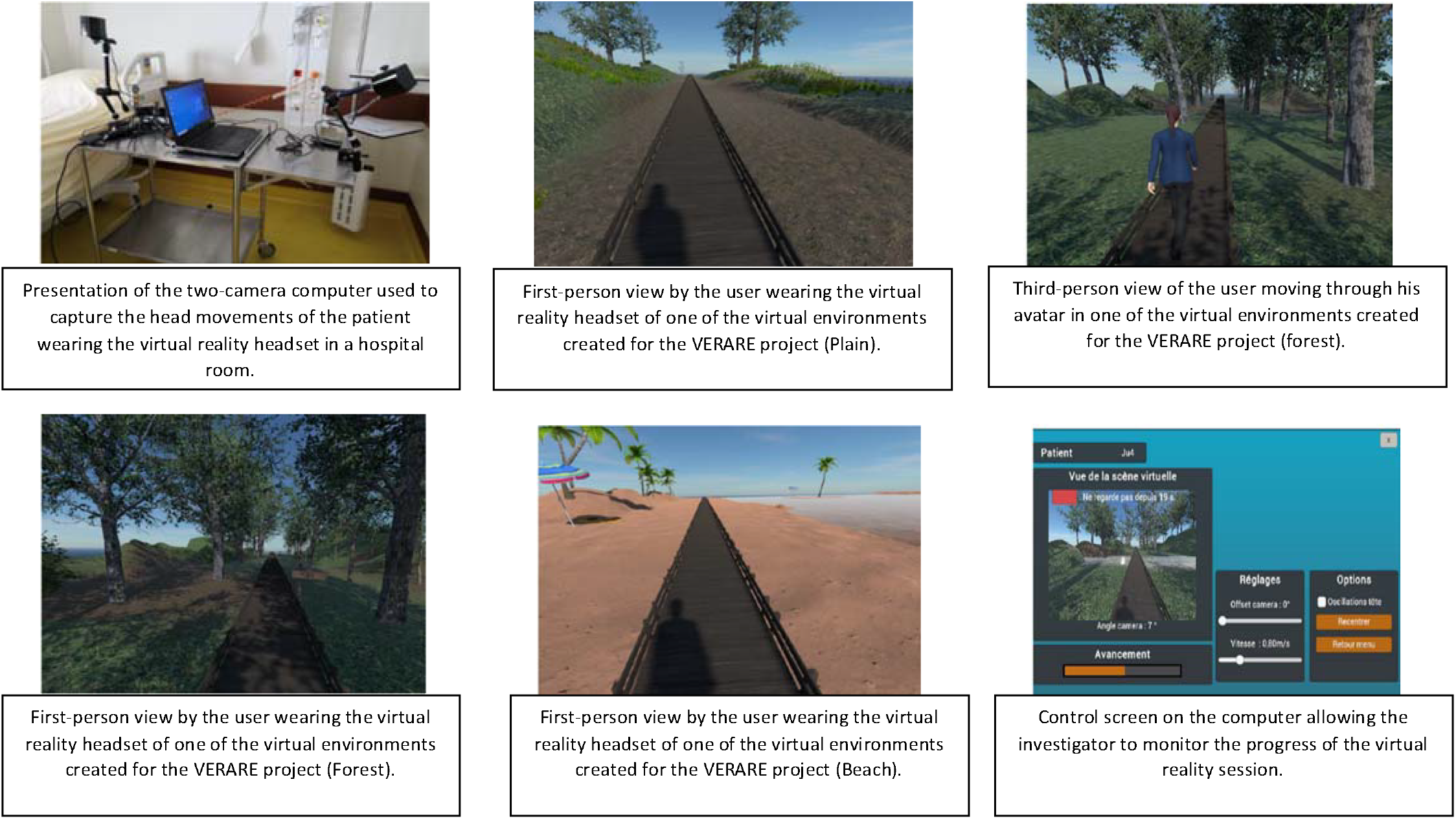

